# Rare gain-of-function regulatory mutations explain the missing heritability of bicuspid aortic valve

**DOI:** 10.1101/2025.02.18.25322302

**Authors:** Artemy Zhigulev, Madeleine Petersson Sjögren, Andrey Buyan, Vladimir Nozdrin, Enikő Lázár, Raphaël Mauron, Karin Lång, Rapolas Spalinskas, Doreen Schwochow, Sailendra Pradhananga, Anders Franco-Cereceda, Joakim Lundeberg, Ivan V. Kulakovskiy, Per Eriksson, Hanna M Björck, Pelin Sahlén

## Abstract

Bicuspid aortic valve (BAV), a prevalent congenital cardiac defect, predisposes patients to severe complications. Despite its high heritability, previously identified protein-coding and common regulatory mutations account for only a small fraction of cases. To address this gap, we investigated the role of rare regulatory mutations. By integrating high-resolution three-dimensional genome organization profiling with whole-genome sequencing, we analyzed sixteen patients with BAV and normal tricuspid aortic valves. Our findings reveal a 1.5-fold enrichment of gain-of-function regulatory mutations in previously implicated genes among BAV patients. Genome-wide, moderately rare mutations (allele frequencies below 3%) were predicted to alter the transcriptome of specific developmental valve mesenchymal cell and fibroblast populations. Expanding the BAV pathway network with newly implicated genes uncovered substantial genetic heterogeneity underlying the disease. These results position rare regulatory mutations as pivotal contributors to missing BAV heritability and emphasize the need for further research of their mechanistic roles.

## Introduction

Bicuspid aortic valve (BAV) is the most common congenital cardiac defect, affecting 0.5 – 2% of the global population [1–3]. Unlike the normal tricuspid aortic valve (TAV), BAV forms with two instead of three leaflets, predisposing individuals to severe cardiovascular complications, including aortic valve stenosis and ascending aortic aneurysm [4]. More than half of BAV patients require surgical interventions, such as valve replacement or repair, during their lifetime [5].

Despite its estimated heritability of up to 90% [6], the genetic etiology of BAV remains largely unresolved [7]. While protein-coding mutations in genes such as *NOTCH1* [8], and *ROBO4* [9] were implicated in familial cases, genome-wide association studies (GWAS) identified only a limited number of common regulatory mutations, including those near *GATA4* [10]. Collectively, these findings explain less than 10% of cases, underscoring the need to investigate rare regulatory mutations – an understudied factor in BAV research and human genetics more broadly [11].

Regulatory mutations modulate gene activity by altering protein-DNA recognition in regulatory regions (promoters and enhancers), thereby disrupting or creating promoter-enhancer interactions [12]. Notably, more than two-thirds of enhancers regulate genes that are not their closest neighbors, necessitating comprehensive mapping of these interactions to uncover disease mechanisms [13]. Emerging evidence suggests that deleterious regulatory interactions active during embryogenesis may leave detectable traces in adult cells [14,15].

Building on this premise, we employed a novel genome-wide integrative approach to uncover the role of rare regulatory mutations in BAV formation. We combined patient-specific promoter-enhancer interactome mapping in aortic endothelial cells (AEC) using promoter capture HiC (HiCap) [16] with whole-genome sequencing (WGS) (Fig. 1). This strategy allowed us to identify causative regulatory interactions and address the missing heritability of BAV, advancing our understanding of its molecular etiology.

**Fig. 1:**
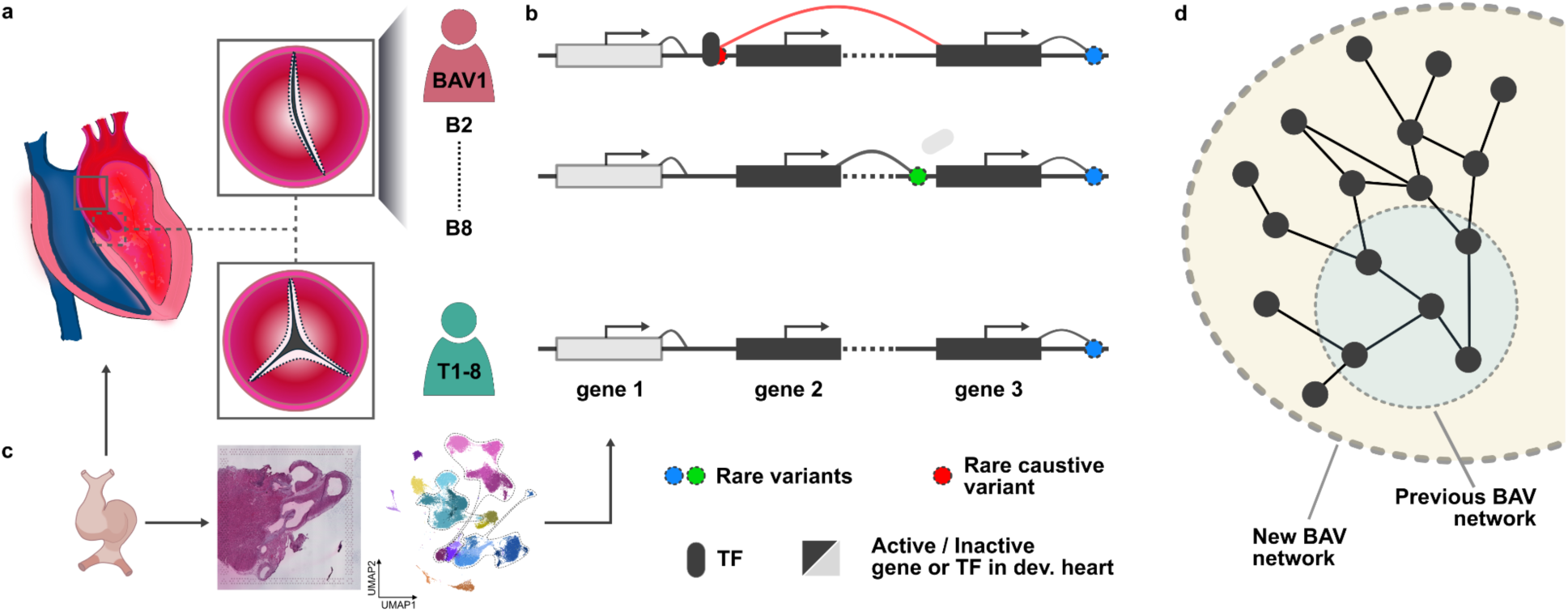
Study overview. **a,** AEC were isolated from the ascending aorta (solid grey square) of adult patients with BAV or TAV (dashed grey square) to map patient-specific promoter-enhancer interactions. **b,** Regulatory interactions were integrated with WGS data from the same patients to identify mutations that create or disrupt interactions. **c,** Developmental heart scRNA-seq and ST datasets were used to prioritize causative interactions active during aortic valve formation, define the MAF range of mutations, and predict affected developmental cell states. **d,** Findings expanded the BAV pathway network, providing an advanced resource for BAV research. AEC, aortic endothelial cells; BAV or B, bicuspid aortic valve; MAF, minor allele frequency; scRNA-seq, single-cell RNA sequencing; ST, spatial transcriptomics; TAV or T, tricuspid aortic valve; TF, transcription factor; WGS, whole-genome sequencing.

## Results

### Genes implicated in aortic valve development are enriched for gain-of-function regulatory mutations in BAV patients

The study included eight BAV and eight TAV patients undergoing elective open-heart surgery at Karolinska University Hospital, all with homogenous clinical profiles (Supplementary Table 1). WGS identified an average of 323,387 ± 18,973 short germline variants, including single nucleotide variants (SNVs) and indels with a minor allele frequency (MAF) below 10% (Supplementary Fig. 1). Structural variants (SVs) were also annotated. Consistent with previous studies, only a few patients exhibited potentially causal protein-coding mutations, including SVs, in disease-relevant *(tier 1)* or general aortic valve *(tier 2)* genes, underscoring the likely contribution of non-coding variants to BAV pathogenesis (Methods, Supplementary Table 2).

Non-coding variants accounted for up to 95% of the identified variation. To prioritize regulatory mutations, we mapped promoter-enhancer interactions in ascending AEC from all patients using HiCap [17]. Ascending AEC serve as a relevant model for studying BAV pathology due to their shared embryonic lineage with aortic valve cells [18]. HiCap targeted 23,111 promoters and 1,123 selected variants (Methods), achieving whole-genome coverage and near single-enhancer resolution (821 bp). In total, we identified 1,264,834 statistically significant promoter-enhancer interactions (Methods, Supplementary Table 3).

To validate the HiCap dataset, we confirmed its ability to capture functional endothelial-specific interactions (Fig. 2a-b). Additionally, HiCap-detected regulatory elements significantly overlapped with H3K27ac ChIP-Seq peaks in human ascending AEC (Methods), as well as with enhancer regions in the TeloHAEC cell line [19], further supporting its capacity to detect endothelial-specific enhancers (Fig. 2c). Notably, we also identified a subset of patient-specific enhancers, highlighting inter-individual variability in regulatory landscapes.

**Fig. 2:**
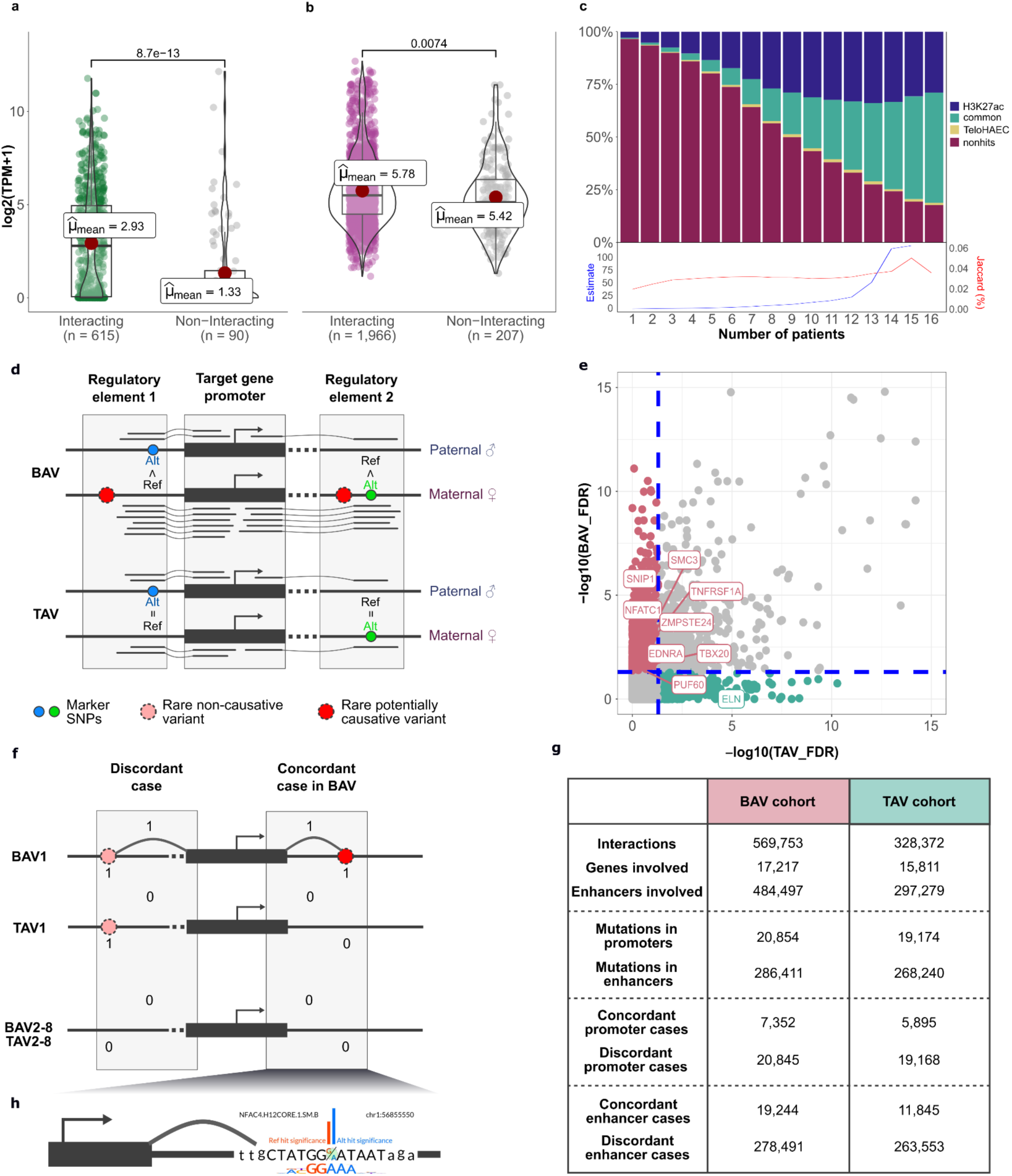
Interactome analysis and filtering strategy. Comparison of expression levels between interacting and non-interacting **(a)** endothelial-specific and **(b)** housekeeping genes. Statistical significance was assessed using a Student’s t-test. **c,** Overlap of HiCap-derived promoter interacting regions with H3K27ac ChIP-Seq peaks from different patients and cumulative TeloHAEC enhancer regions. The estimate of the difference between HiCap and H3K27ac regions is in blue, while the Jaccard similarity index is in red. **d,** Schematic of allele-specific interaction analysis. Arches represent chimeric reads spanning the ligation sites **e,** Per-gene aggregation of allele-specific interaction enrichments in BAV and TAV cohorts. FDR thresholds of 0.05 (blue dashed lines) are shown for TAV and BAV values. **f,** Representation of concordant and discordant interactions. **g,** Numerical overview of the filtering strategy. **h,** Schematic of TF binding alterations caused by mutations.. BAV, bicuspid aortic valve; HiCap, sequence capture HiC; TAV, tricuspid aortic valve; TPM, transcript per million.

Rare regulatory mutations are typically heterozygous, affecting promoter-enhancer interactions on a single allele (Fig. 2d). Direct allelic imbalance analysis of HiCap data, conducted without incorporating WGS, revealed significant signals in multiple *tier 1* (*EDNRA, PUF60, SNIP1, SMC3*) and *tier 2* (*NFATC1, TBX20, TNFRSF1A, ZMPSTE24*) genes specific to BAV patients (Fig. 2e, Supplementary Table 4). Notably, *ELN* was the only gene exhibiting allelic imbalance signals specific to TAV patients. Overall, genes with allelicly-imbalanced interactions were overrepresented in *tier 1* and *tier 2* gene sets in the BAV cohort (p = 0.03537, OR = 8.470844, two-tailed Fisher’s exact test). These findings support the integration of HiCap with WGS at the next step as an effective strategy for uncovering the regulatory effects of mutations.

To identify causative promoter-enhancer interactions, we annotated interacting regulatory regions with 53,784 promoter and 745,382 enhancer variants detected in the patients (MAF < 10%). Cases where interactions were observed only in patients carrying a variant in the interacting promoter or enhancer were classified as *concordant*, assuming the variant affected the interaction (Fig. 2f). Mismatched cases were designated as *discordant*. We further categorized concordant and discordant cases by BAV and TAV cohorts (Fig. 2g). Downstream analyses were conducted in parallel for BAV- and TAV-cohort specific interactions, using discordant cases and the TAV cohort as internal controls. Notably, BAV-specific concordant interactions were more prevalent in *tier 1* and *tier 2* genes than in housekeeping genes compared to TAV-specific cases (p = 1.858e-15, OR = 1.536895, two-tailed Fisher’s exact test), emphasizing their predictive value for BAV pathogenesis. In contrast, discordant interactions showed minimal enrichment in the TAV cohort (p < 3.478e-08, OR = 0.9115922, two-tailed Fisher’s exact test).

To refine our analyses, we focused on cohort-specific concordant mutations predicted to alter transcription factor (TF) binding (Fig. 3h, Supplementary Table 5). Using a non-redundant binding motif subset from HOCOMOCO v12 [20], we identified the overrepresentation of NFAT-related TF family binding site alterations in BAV patients compared to TAV individuals (FDR < 1.19e-20, OR = 1.42, two-tailed Fisher’s exact test). Strikingly, only two disrupted concordant interactions were linked to altered TF binding. Instead, most cases involved the establishment of novel promoter-enhancer interactions, suggesting a gain-of-function mechanism that may play a key role in BAV pathogenesis.

**Fig. 3:**
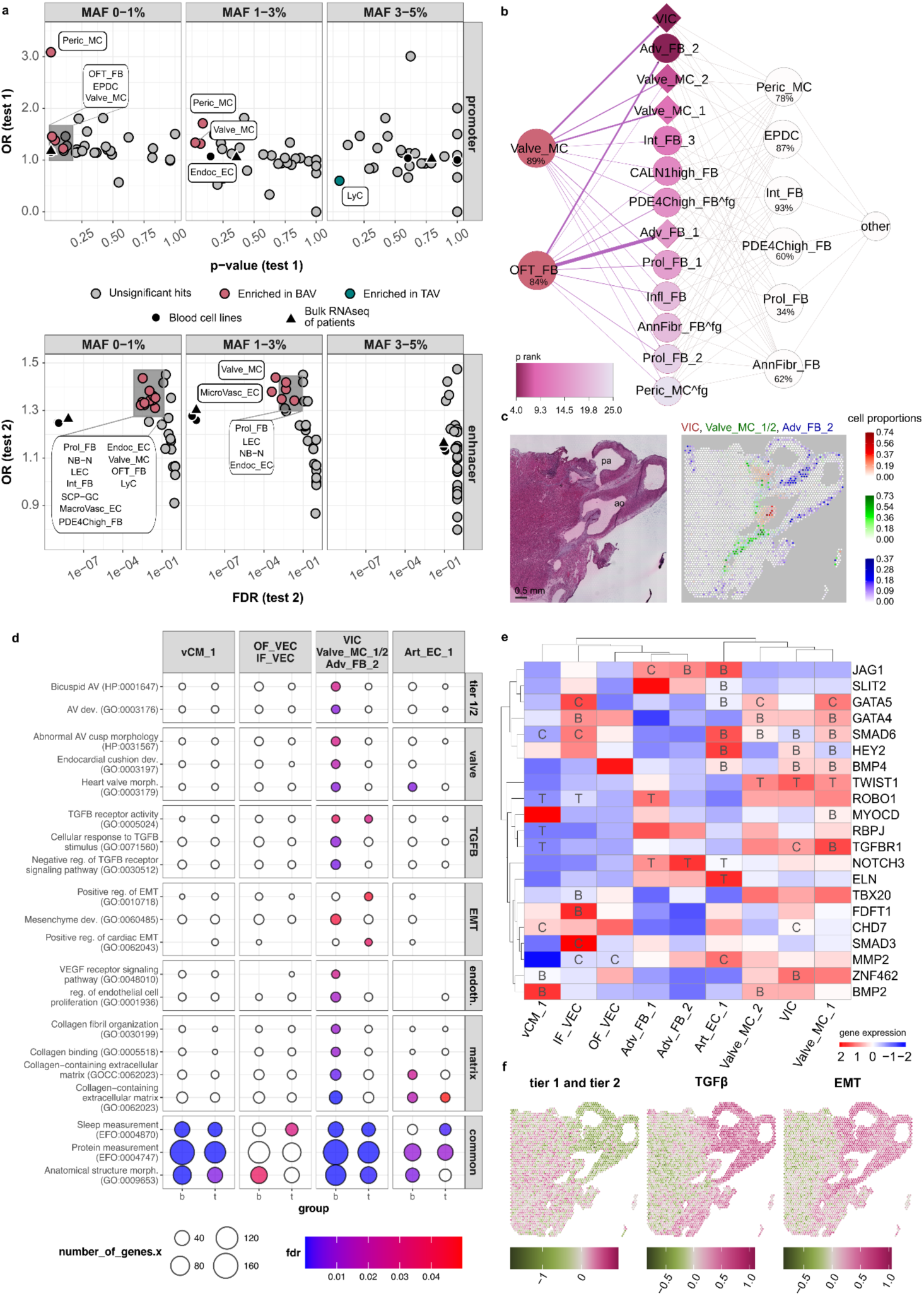
Mechanistic insights into BAV etiology. **a,** Impact of promoter and enhancer concordant mutations across different MAF ranges on developmental cell types (two-tailed Fisher’s exact test). Different tests were applied for promoter (test 1) and enhancer (test 2) mutations. DE genes between the cell types were included (Wilcoxon rank sum test). **b,** Prioritization of affected cell states based on top 2500 expressed genes. P rank corresponds to test 1 / test 2 rankings. Purple edges indicate the proportion of cell states derived from Valve_MC and OFT_FB (red), while contributions from other cell types (white) are represented with grey edges. Percentages in cell type nodes denote the fraction of cells lacking fine-grained annotation. Cell type annotation numbers correspond to different cell states. **c,** Spatial mapping of affected cell states (cell state proportion per Visium spot) VIC (red), Valve_MC_1&Valve_MC_2 (green) and Adv_FB_2 (blue). The proportions of Valve_MC_1&Valve_MC_2 were summed up. The three results are represented on a single figure for better understanding of their spatial relationships. **d,** Gene set enrichment analysis of concordant genes in spatially prioritized cell states. DE genes were used. **e,** Gene-level analysis of tier 1 and tier 2 genes affected by concordant mutations in BAV (B), TAV (T), and shared (common - C) cohorts, linked to overall expression levels. Hierarchical clustering was performed based on the B (3), T (-3), C (1), NA (0) matrix. DE genes were analyzed. **f,** Spatial mapping of concordantly affected genes within key gene sets. Adv_FB_1/2, adventitial fibroblast states 1 and 2; AnnFibr_FB, annulus fibrosus fibroblasts; BAV, bicuspid aortic valve; CALN1high_FB, CALN1-enriched fibroblast population; DE, differentially expressed; Endoc_EC, endocardial cells; EPDC, epicardium-derived progenitor cells; fg, fine-grained; Infl_FB, inflammatory mediator-enriched fibroblasts; Int_FB, interstitial fibroblasts; LEC, lymphatic endothelial cells; LyC, lymphoid cells; MacroVasc_EC, macrovascular endothelial cells; MAF, minor allele frequency; MicroVasc_EC, microvascular endothelial cells; NB-N, neuroblasts and neurons; OFT_FB, fibroblasts around the outflow tract and developing great arteries; PDE4Chigh_FB, PDE4C-enriched fibroblasts; Peric_MC, cardiac pericyte; Prol_FB, proliferating fibroblasts; scRNA-seq, single-cell RNA sequencing; SCP-GC, Schwann cell progenitors and glial cells; ST, spatial transcriptomics; TAV, tricuspid aortic valve; Valve_MC, cardiac valve-related mesenchymal cells; VIC, valve interstitial cells.

### Rare regulatory mutations in BAV patients alter the transcriptome of fetal heart mesenchymal cells and fibroblasts

To determine the MAF range of causative concordant regulatory mutations and accordingly predict their transcriptomic impact on heart cell types, we analyzed a developmental single-cell RNA sequencing (scRNA-seq) dataset spanning the critical developmental window of aortic valve morphogenesis (5.5–14 weeks post-conception) [21]. This dataset integrates scRNA-seq with spatial transcriptomics (ST), profiling 31 coarse-grained cell types and 72 fine-grained cell states, providing a comprehensive framework for investigating developmental heart abnormalities.

We categorized concordant and discordant mutations into three MAF ranges: rare (MAFa, 0–1%), moderately rare (MAFb, 1–3%), and common (MAFc, 3–5%). Promoter and enhancer mutations were analyzed separately using distinct statistical tests to account for sample size differences (Methods). In the BAV cohort, transcriptomes of cardiac valve-associated mesenchymal cells (Valve_MC) and fibroblasts near the outflow tract and great arteries (OFT_FB) were the most affected (p < 0.01, two-tailed Fisher’s exact test) cell types by concordant promoter and enhancer mutations within the MAFa/MAFb ranges compared to the TAV cohort (Fig. 3a). In contrast, the TAV cohort exhibited significantly different transcriptomic effects only by MAFc promoter mutations in lymphoid cells, a cell type without known relevance to valve development. These results highlight the importance of rare and moderately rare mutations in BAV.

The fine-grained resolution of the single-cell dataset provided deeper insights into the cellular impact of regulatory mutations. Within the MAFa/MAFb ranges, the most strongly implicated (FDR < 0.05, two-tailed Fisher’s exact test) fine-grained cell states in the BAV cohort included valve interstitial cells (VIC), valve mesenchymal cell states 1 and 2 (all three derived from Valve_MC), and adventitial fibroblast state 2 (derived from OFT_FB) (Methods, Fig. 3b). These cell states collectively belong to a mesenchymal-fibroblast subset (MC/FB). ST data confirmed their spatial proximity to the aortic valve region, underscoring their relevance to BAV pathogenesis (Fig. 3c).

We then performed gene set enrichment analyses of concordant and discordant genes altering the transcriptomes of prioritized MC/FB cell states and other spatially relevant cell states. These included valve endothelial cells on the inflow (IF_VEC) and outflow (OF_VEC) sides of the valves, as well as arterial endothelial cells (Art_EC_1) from the endothelial subset (EN), and ventricular cardiomyocytes vCM_1 from the cardiomyocyte subset (CM) [21]. Concordant BAV cases were significantly enriched for *tier 1* and *tier 2* genes and pathways related to valve development, transforming growth factor beta (TGFβ) signaling, epithelial-to-mesenchymal transition (EMT), and extracellular matrix (ECM) formation, predominantly in the MC/FB fine-grained cell states (Fig. 3d). In contrast, discordant cases showed nonspecific enrichment patterns (Supplementary Fig. 2).

While MC/FB cell states exhibited strong and consistent enrichments, EN cell states showed limited signals, and CM cells were minimally affected by concordant mutations. A heatmap of *tier 1* and *tier 2* genes affected by concordant mutations in BAV and TAV cohorts reinforced this trend (Fig. 3e). Further spatial analysis of the affected genes within the highlighted pathways revealed distinct expression patterns between the BAV and TAV cohorts (Fig. 3f). Notably, concordantly affected *tier 1* and *tier 2* genes did not show specific spatial enrichment unique to the BAV cohort compared to those in the TGFβ and EMT pathway.

By defining relevant MAF ranges and affected cell states, we refined our dataset of concordant interactions, yielding a final list of 258 BAV-specific and 161 TAV-specific genes (Supplementary Table 6).

### Newly implicated genes in BAV etiology highlight its high heterogeneity

To investigate the molecular mechanisms underlying BAV, we constructed protein-protein interaction (PPI) networks for prioritized concordantly affected genes specific to the BAV or TAV cohorts (Methods). The BAV-specific network (nodes = 266, edges = 189, average node degree = 1.42, PPI enrichment p < 1.0e-16) exhibited more interactions than the TAV-specific network (nodes = 164, edges = 59, average node degree = 0.72, PPI enrichment p = 6.08e-07) (Fig. 4a-b). The BAV network included significantly more genes than the *tier 1* and *tier 2* sets, highlighting key pathway interactions, including collagen biosynthesis and vascular endothelial growth factor signaling, thus providing novel insights into BAV genetics.

**Fig. 4:**
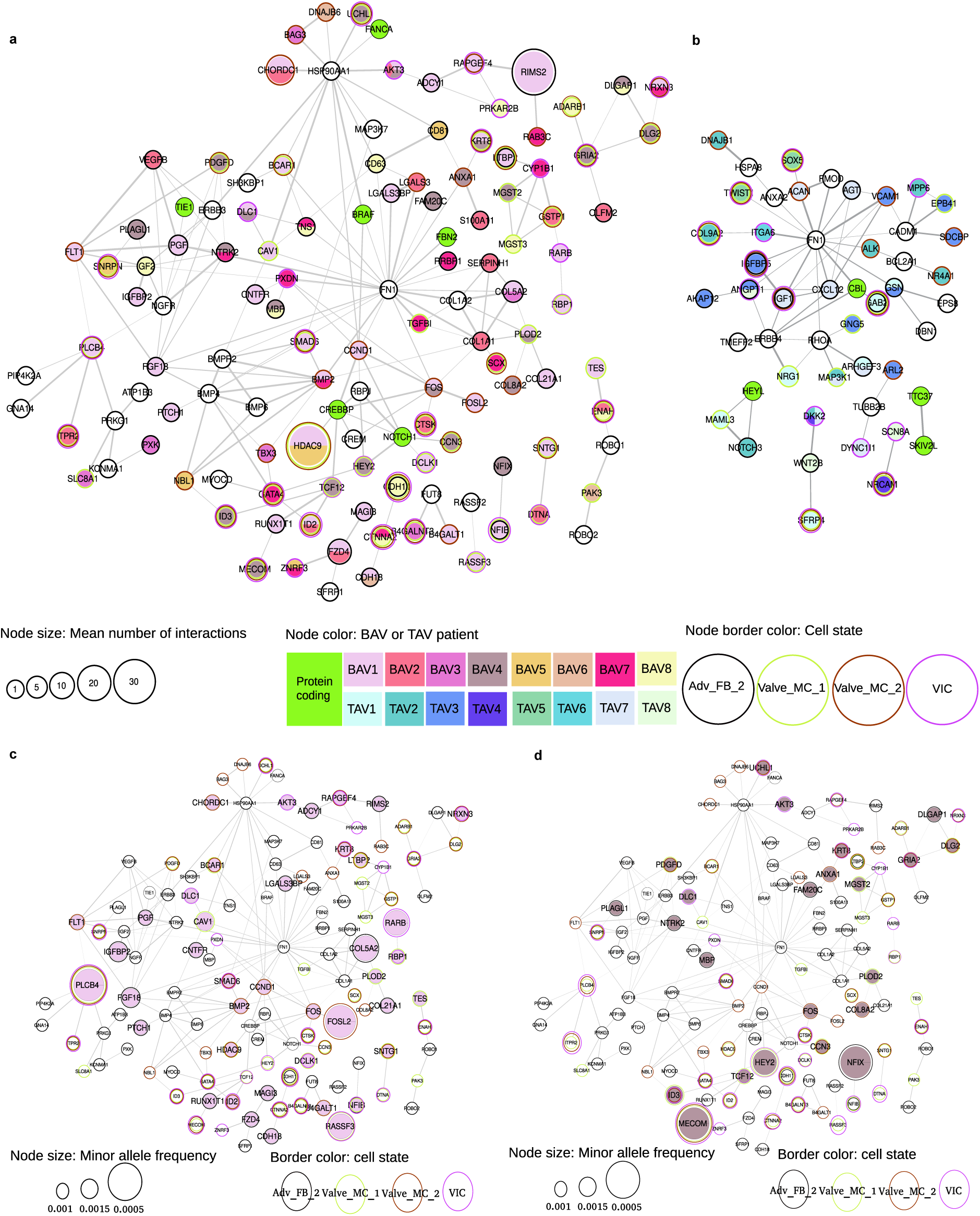
Extended BAV pathway network and single-patient analysis. PPI networks of **(a)** BAV-specific and **(b)** TAV-specific concordant genes and protein-coding mutations. Isolated nodes are omitted for clarity. Node colors represent individual patients. Node size reflects the mean number of interactions. Edge thickness indicates interaction confidence scores. Aditional nodes with no mutations (white) were directly connected to the prioritized mutated genes for better network integrity. Single-patient PPI networks of **(c)** BAV1 and **(d)** BAV4, illustrating patient-specific genetic landscapes. Node size reflects MAF of underlying variants. BAV, bicuspid aortic valve; MAF, minor allele frequency; PPI, protein-protein interaction; TAV, tricuspid aortic valve.

To further explore the heterogeneity of BAV etiology, we constructed single-patient gene networks. Notably, six BAV patients carrying high-to-moderate impact protein-coding short mutations or SVs exhibited relatively smaller regulatory networks (Supplementary Fig. 3-5). Analysis of the two most extensive individual networks within the BAV cohort revealed a complex genetic landscape underlying the disease (Fig. 4c–d). The colocalization of rare and more common mutations, alongside protein-coding alterations, suggests the existence of a multilayered regulatory mechanism contributing to BAV formation.

Overall, our findings underscore the contribution of rare regulatory mutations to BAV formation and highlight the intricate genetic interplay between different mutation classes that may underlie its etiology.

## Discussion

This study presents the first integrative whole-genome interactome analysis of BAV and TAV patients, combining promoter-enhancer interaction mapping from HiCap data with WGS. Unlike previous GWAS that primarily focused on common regulatory mutations and linked them to the nearest genes, our approach allows for the identification of all regulatory mutations, regardless of MAF, and their precise mapping to target genes. As a result, we uncovered a ignificant enrichment of gain-of-function regulatory mutations in aortic valve developmental genes, specifically within the BAV cohort. Additionally, our human interactome dataset provides a valuable resource for identifying other disease-related regulatory changes [22–25].

By integrating our findings with a human developmental heart scRNA-seq and ST data, we explored the molecular mechanisms underlying BAV pathogenesis. Our results demonstrate that rare and moderately rare regulatory mutations (MAF < 3%) significantly reshape the transcriptomic landscape of developmental valve mesenchymal cells and outflow tract fibroblasts. One particularly notable finding is the systematic role of the NFAT-related TF family [26,27]. While *NFATC1* exhibited allele-specific interactions, binding site alterations across the entire NFAT TF family were overrepresented in BAV patients.

Among valve-related developmental cell states, rare regulatory mutations prioritized from adult endothlial cells were more frequently associated with expressed genes in mesenchymal cells and fibroblasts than in endothelial or cardiomyocyte subsets. However, whether valve endothelial and interstitial cells or ascending aortic smooth muscle cells samples exhibit distinct regulatory signals remains an open question, emphasizing a key area for future investigation.

A particularly intriguing finding is the genome-wide retention of developmental regulatory interactions in adult patients [14,15]. This indicates that certain regulatory interactions observed in adult endothelial cells can be traced back to developmental events in cardiac fibroblasts and mesenchymal cells, despite differences in their gene expression programs. These retained interactions appear to be actuated by a specific set of trans-acting factors exclusively during development.

Furthermore, clustering of cell states based on regulatory mutation patterns in aortic valve development genes revealed that Art_EC_1, the endothelial cell state with the closest transcriptomic profile to adult endothelial cells, showed greater similarity to mesenchymal cells and fibroblasts than to other endothelial populations. This finding highlights a potential lineage-passing mechanism of altered developmental interactions. However, further research is required to explore this phenomenon in detail.

Our study underscores the critical role of rare regulatory mutations in BAV pathology and heritability, particularly their influence on developmental mesenchymal and fibroblast populations involved in aortic valve formation. By elucidating the genetic etiology of BAV, this work establishes a foundation for future research into the dynamic interplay between regulatory mutations and cell-type-specific transcriptomic alterations during development and disease progression.

## Methods

### Patient cohorts

In this study, we included eight BAV and eight TAV individuals undergoing elective open-heart surgery for ascending aortic dilatation at the Cardiothoracic Surgery Unit, Karolinska University Hospital, Stockholm, Sweden. None of the individuals had significant coronary artery disease, based on coronary angiography (The ASAP/DAVAACA study). The study was approved by the Human Research Ethics Committee at Karolinska Institutet (application number 2006/784-31/1 and 2012/1633-31/4), Stockholm, Sweden; written informed consent was obtained from all the individuals according to the Declaration of Helsinki, and methods were carried out following relevant guidelines.

### Whole genome sequencing and variant calling

Patient blood samples were collected at baseline before treatment started. The Blood & Cell Culture DNA Midi Kit Kit (Qiagen) was used according to the manufacturer’s protocol to extract DNA from samples. Sequencing libraries were then prepared with the KAPA HTP Library Preparation Kit (Kapa Biosystems) with PCR-free workflow, according to the manufacturer’s protocol. Samples were sequenced on NovaSeq6000 (NovaSeq Control Software 1.7.5/RTA v3.4.4) with a 101nt(Read1)-8nt(Index1)-8nt(Index2)-101nt(Read2) setup using ‘NovaSeqXp’ workflow in ‘S4’ mode flowcell. The Bcl to FastQ conversion was performed using bcl2fastq_v2.20.0.422 from the CASAVA software suite. The quality scale used is Sanger / phred33 / Illumina 1.8+.

FastQ files were then processed using the nf-core/sarek/2.7.1 pipeline [28–30]. Briefly, sequences were aligned to GRCh38 (BWA) [31] and preprocessed based on GATK4 Best Practices (GATK/4.1.7.0). SNPs and small indels were called using GATK HaplotypeCaller [32], recalibrated according to GATK Variant Quality Score Recalibration algorithm, and annotated with snpEff/4.3t [33]. Non-coding effects included the following annotations: intron_variant, intergenic_region, downstream_gene_variant, upstream_gene_variant, intragenic_variant, TF_binding_site_variant, TFBS_ablation. SweGen/20190204 [34] was used for initial filtering, multi-allelic variants were excluded. Structural Variants were called using Manta [35] (nf-core/sarek pipeline v3.1.2, Manta v. 1.6.0) and annotated with ensemblVEP/106.1 [36] and snpEff/5.1d [33].

### *Tier 1* and *tier 2* gene sets establishment

*Tier 1* gene set, comprising genes previously associated with BAV (HP:0001647, DOID:0080332) and *tier 2* gene set, which included genes associated with more general aortic valve terms (GO:0003176, HP:0001646), were built based on the STRINGdb v12 [37] (R package v2.12.1) (species=9606, score_threshold=200, network_type=“full”).

### Analysis of protein coding mutations

High and moderate impact mutations were included in the analysis. Short variants were additionally annotated with GnomAD v4 [38] with the overall MAFmax < 0.001. For moderate impact short variants changes in hydropathy, charge and polarity of amino acids were used for prioritization. All high and moderate impact SVs were included.

### Aortic biopsy collection

Aortic biopsies were collected directly in the operating theatre and washed in PBS containing Calcium chloride and Magnesium chloride. The medial layer was separated from the adventitia using adventectomy and subjected to enzymatic digestion using a solution of 1mg/mL collagenase A (Roche, 11088793001) in dispase 1U/mL (Stemcell Technologies, 07923) for 20 to 25 min at 37°C, with regular gentle rocking of the Petri dish. After incubation, the endothelial side of the tissue was carefully scraped 5 to 7 times using a sterile scalpel. The collagenase solution containing endothelial cells was collected in a tube, and the tissue was rinsed a couple of times with PBS and collected in the same tube. The solution was then strained using a 100μm cell strainer and centrifuged at 400g for 5 min. Finally, the pellet was resuspended in 2.5mL of EBM-2 basal medium supplemented with EGM-2 BulletKit (Lonza, CC-3162) and dispensed in a 12.5cm2 flask previously coated with 0.2% bovine gelatin type B (Sigma, G1393). The next day, cells were gently washed twice with PBS, and a fresh endothelial cell growth medium was added. Hereafter, the medium was replaced every 2 to 3 days. Upon confluence, cells were transferred to a 75cm2 flask and frozen at subconfluency in a solution of 90% FBS +10% DMSO or used no later than P4. The primary endothelial cells were purified with CD31 beads (Miltenyi Biotec, #130-091-935) in MS columns (Miltenyi Biotec, #130-042-201) before use.

### HiCap and analysis

We designed 49,927 probes targeting 23,111 promoters (of 19,469 genes), 1,123 GWAS variants associated with cardiovascular disease and lipid traits and 2,408 negative controls regions with no known regulatory activity (Supplementary Table 7).

HiCap was performed on 2,5 million cells as previously explained [17]. This method was developed by us [16] and others and provides high-resolution interaction data over genomic regions by hybridizing Hi-C material to probes targeting certain regions of interest, enabling the study of individual promoter-enhancer interactions. Briefly, the method starts with cross-linking DNA-protein-DNA complexes with formaldehyde, followed by roughly cutting DNA across the genome into ∼700 bp fragments using restriction endonucleases. Spatially close fragments are then ligated before capturing promoter-enhancer sequences using probes located in known genes’ promoters or probes containing selected SNVs associated with cardiovascular disease and lipid traits. Captured libraries were sequenced using HiSeq 2500 (Illumina) with HiSeq Rapid SBS Kit v2 chemistry and a 1×80 setup at Science for Life Laboratory (SciLifeLab, Stockholm, Sweden).

Lastly, sequencing data was analyzed for significant interactions. We mapped HiCap libraries using bwa-mem2 (version 2.2.1-20211213-edc703f) [31] and PairTools (version 1.0.2) [39]. In total, we processed 15.56 billion reads, of which 11.25 billion were uniquely mapped. Read statistics are found in table (Supplementary Table 8). HiCapTools [40] was used to call interactions in all samples. We required five or more pairs supporting each interaction and Bonferroni-adjusted P-value < 0.05. We further filtered out trans interactions, long-range interactions with an interaction distance higher or equal to 5Mb, and PIRs/DpnII fragments with lengths higher or equal to 7,500 bp.

### Bulk RNA-Seq and analysis

Total RNA from 16 samples was sequenced in two batches (6 and 10 samples correspondingly). In both cases, rRNA was depleted using RiboCop Kit (Corall) and final libraries were prepared using Total RNA-Seq Library Prep Kit (Corall). Samples were sequenced on NovaSeq6000 (NovaSeq Control Software 1.7.5/RTA v3.4.4) with a 51nt(Read1)-8nt(Index1)-8nt(Index2)-51nt(Read2) setup using ‘NovaSeqXp’ workflow in ‘SP’ mode flowcell. The Bcl to FastQ conversion was performed using bcl2fastq_v2.20.0.422 from the CASAVA software suite. The quality scale used is Sanger / phred33 / Illumina 1.8+. FASTQ files were processed using nf-core/rnaseq pipeline version 3.5 within nf-core framework [30].

For the downstream analysis, six samples from the first experimental batch were excluded due to a 10-fold difference in sequencing coverage compared to the second batch. Additionally, two outliers from the second batch, BAV2 and TAV3 were removed from the analysis. DE genes were called using the edgeR package [41] based on raw gene counts (Supplementary Table 9, Supplementary Fig. 6).

### ChIP-Seq and peak calling

Human ascending AEC from four patients were cross-linked with 1% formaldehyde for 10 min. Around 5 million cells were used for each experiment and each experiment was performed in duplicates. All steps were performed at 4°C unless otherwise indicated. Cells were lysed in swelling buffers (100 mM Tris at pH 7.5, 10 mM KOAc, 15 mM MgOAc, 1% igepal, PIC) for 10 min followed by Dounce homogenization. Nuclei were pelleted at 4000 rpm for 5 min and lysed in a modified RIPA buffer (PBS with 1% NP-40, 0.5% sodium deoxycholate, 0.1% SDS, 1 mM EDTA, PIC) for 10 min. Chromatin was sonicated to 150–300 bp using a Diagenode bioruptor. Insoluble material was removed by centrifugation at 13,000 rpm for 10 min followed by preclear with StaphA cells for 15 min. One microgram of primary antibody/5 million cells (Diagenode, C15410196-10) was incubated with precleared chromatin for 16 h. Anti-rabbit secondary antibodies (Millipore) were added for 1 h. Ten microliters of StaphA cells/5 million cells were added for 15 min at room temperature. StaphA cells were washed twice with dialysis buffer (50 mM Tris at pH 8, 2 mM EDTA, 0.2% sarkosyl) and four times with immunoprecipitation wash buffer (100 mM Tris at pH 8, 500 mM LiCl, 1% NP-40, 1% sodium deoxycholate). Chromatin was eluted off StaphA cells in 1% SDS and 50 mM NaHCO3. For re-ChIP, eluted chromatin was diluted 10-fold in modified RIPA without SDS, and a second primary antibody was added overnight. For ChIP-seq and ChIP-qPCR, 200 mM NaCl was added, and cross-links were reversed for 16 h at 67°C. DNA was purified using a PCR purification kit (Qiagen), eluting in 50 μL of water. One microliter of ChIP DNA was used for qPCR. For ChIP-seq, libraries were prepared using the Mondrian (NuGen) and size-selected using Pippin Prep (Sage Science). Libraries were sequenced with 1 Å∼ 50 base pair read on the NextSeq 500 platform (Illumina Inc). FastQ files were then processed using the nf-core/chipseq/1.2.2 pipeline [30] (Supplementary Table 10).

### Allele-specific analysis of the HiCap data

Variant calling from pre-made HiCap read alignments was performed using bcftools (v.1.21) [42] mpileup with --redo-BAQ --adjust-MQ 50 --gap-frac 0.05 --max-depth 10000 -A parameters to include read pairs that map far apart and bcftools call with --keep-alts --multiallelic-caller. The resulting sites were split into biallelic records using bcftools norm with --check-ref x -m - and filtered with bcftools filter -i “QUAL>=10 & FORMAT/GQ>=20 & FORMAT/DP>=10” -- SnpGap 3 --IndelGap 10 leaving only high-quality calls covered by 10 or more reads. Next, bcftools view -i ‘GT=“alt”’ --trim-alt-alleles followed by view -m2 -M2 -v snps was used to leave only biallelic SNVs annotated with bcftools +fill-tags -- -t all. Finally, filters for heterozygous variants located on the reference chromosomes with genotype quality ≥ 50, depth ≥ 10, and allelic counts ≥ 5 for each allele were applied to each individual separately with bcftools view -s <INDIVIDUAL= -i ‘FORMAT/AD[0:0]>4 & FORMAT/AD[0:1]>4>=“het”’ -e ‘GT=“mis” | FMT/DP<10 | FMT/GQ<50’. The allelic imbalance was assessed with MIXALIME (v.2.25.2) [43] using the beta negative binomial model and individual-level adaptive coverage filters enabled by MIXALIME combine --adaptive-min-cover --group <INDIVIDUAL=. Variants were then annotated with the significantly interacting enhancer and promoter regions derived from HiCap.

MIXALIME provides one-tailed P-values. As we were not focusing on the direction of the allelic imbalance, the minimal P-values (reflecting the imbalance towards either Ref or Alt allele) were doubled to mimic the significance of a two-tailed test. The gene-level P-values were obtained by combining P-values for all SNPs annotated with a particular gene using the logit method [44] implemented in the metap package (v.1.8). P-values of the single variant-per-gene were left as is. Next, the gene-level P-values were re-combined across individuals using the same logitp function from metap followed by Benjamini–Hochberg FDR correction for the number of genes.

### Motif annotation of the single-nucleotide variants

522 PWMs of the highest A-B-C quality from the HOCOMOCO v12 non-redundant set of 523 PWMs [20] were used for motif annotation of the single-nucleotide variants. To this end, we employed PERFECTOS-APE v3.0.6 [45] (https://github.com/autosome-ru/macro-perfectos-ape). Valid motif hits with the P-value ≤ 0.0005 (default threshold) were used to estimate the variant impact, the motif P-value log_2_ fold change between the reference and the alternative alleles.

### scRNA-seq cell type enrichment *test 1* and *test 2*

We performed two statistical tests to assess the impact of regulatory mutations. Test 1 was applied to promoter mutations: using a two-tailed Fisher’s exact test, we compared the proportion of concordant genes between the BAV and TAV cohorts. Test 2 focused on enhancer mutations: similarly, a two-tailed Fisher’s exact test was used to evaluate the proportions of both concordantly and discordantly affected genes across the cohorts. In both tests, target genes and TFs were prefiltered based on their expression levels or differential expression status.

### Protein-Protein Interaction Network Construction and Visualization

BAV and TAV specific protein-protein interaction networks were generated using STRINGdb v12 [37]. For each network, BAV- and TAV-specific genes, along with protein-coding genes, were used as input. The following settings were applied in STRING: - full STRING network, active interaction sources - text mining, experiments and databases, confidence threshold - 0.650, and one shell of interactors with a maximum of one interactor. Disconnected nodes were removed to improve interpretability. Resulting networks were visualized using Cytoscape v3.10.3 [46].

## Data Availability

The sensitive sequencing datasets are not publicly deposited but are available from the corresponding author upon reasonable request. Intermediate anonymized data/results required to replicate the analysis will be shared through Supplementary Tables upon publication.

## Code Availability

Custom codes used in this study will be shared through GitHub upon publication.

## Acknowledgements

The authors acknowledge support from the National Genomics Infrastructure in Stockholm funded by Science for Life Laboratory, the Knut and Alice Wallenberg Foundation, and the Swedish Research Council, and SNIC/Uppsala Multidisciplinary Center for Advanced Computational Science for assistance with massively parallel sequencing and access to the UPPMAX computational infrastructure. P.S. was supported by the European Union’s Horizon 2020 Research and Innovation Programme under the Marie Sklodowska-Curie Grant Agreement No 860002. The information contained in this study reflects only the authors’ views. The European Commission and the European Research Executive Agency do not accept responsibility for any use made of the information contained therein.

## Author Contributions

Conceptualization: P.S., A.Z.

Methodology: P.S., A.Z., I.V.K., E.L.

Software: A.Z., A.B., V.N., M.P.S., S.P.

Validation: E.L., R.M., D.S.

Formal analysis: A.Z., A.B., V.N., S.P., M.P.S.

Investigation: A.Z., R.S., K.L.

Resources: A.F-C., P.S., P.E., H.M.B., J.L.

Data curation: A.Z., P.S.

Writing – original draft: A.Z.

Writing – review and editing: A.Z., P.S., H.M.B., P.E., I.V.K., E.L., M.P.S., A.B., V.N., R.M., K.L.

Visualization: A.Z., M. P. S., R.M., A.B., V.N.

Supervision: P.S., I.V.K., H.M.B., P.E.

Project administration: A.Z., P.S.

Funding acquisition: P.S., P.E., H.M.B.

## Supplementary information

**Fig. S1:**
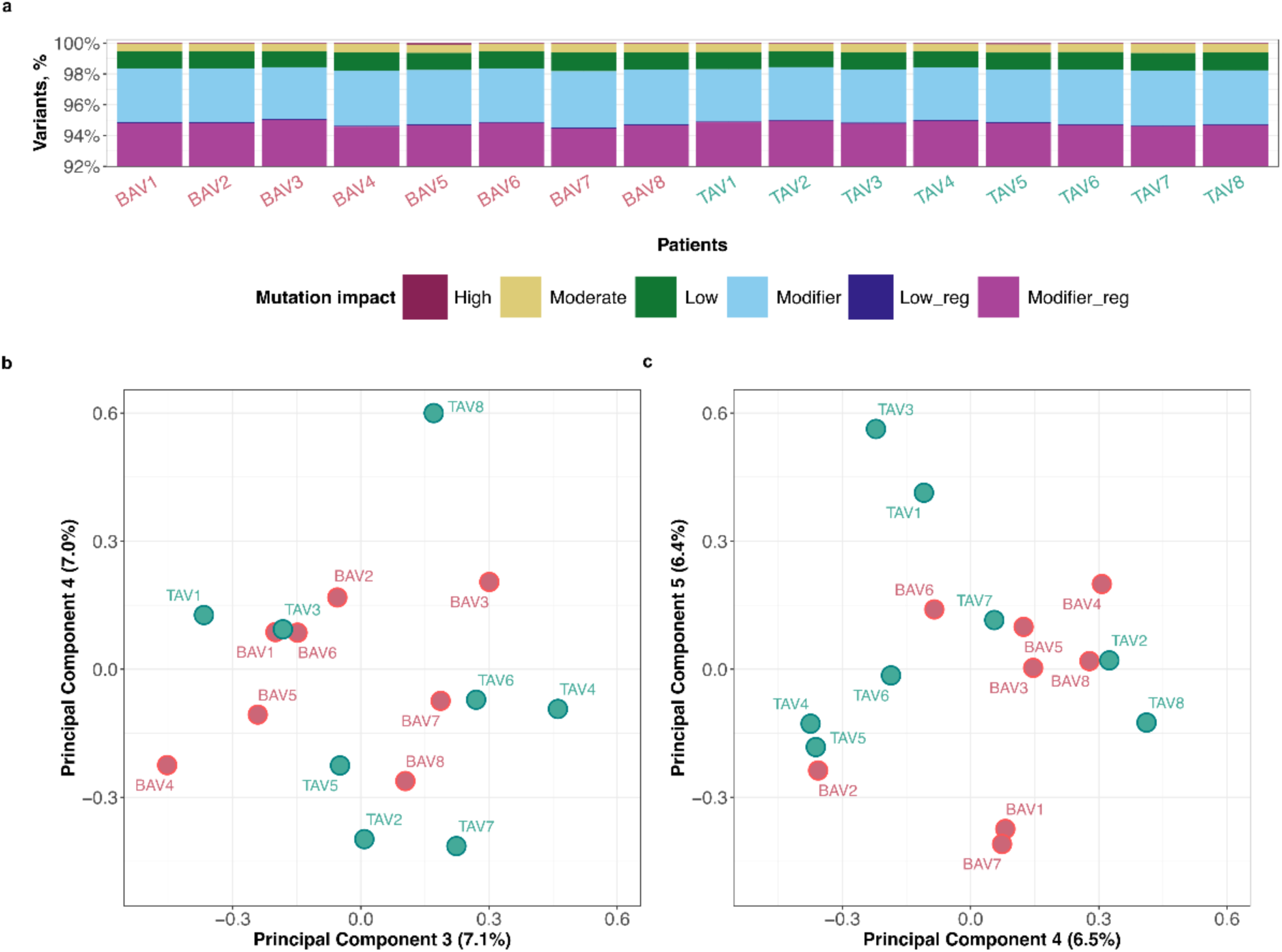
WGS analysis. **a,** Classification of short germline variants by impact and their coding or non-coding/regulatory status. **b,** PCA plot of high-impact protein-coding mutations, showing no distinct clustering between TAV and BAV groups. **c,** PCA plot of non-coding variants, similarly showing no distinct separation between TAV and BAV cohorts.

**Fig. S2:**
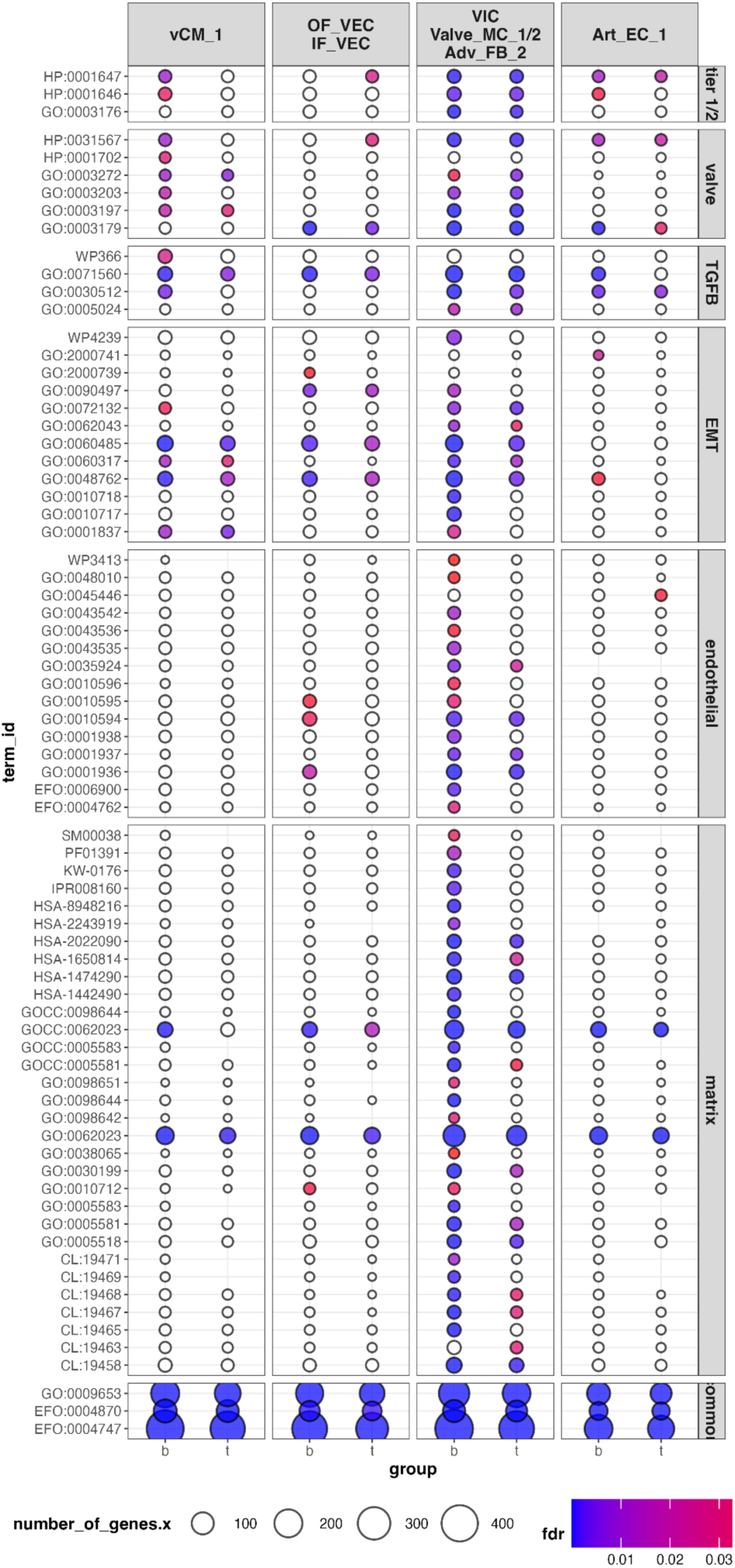
Gene set enrichment analysis of discordant cases. Gene set enrichment analysis of discordant genes across spatially relevant cell states.

**Fig. S3:**
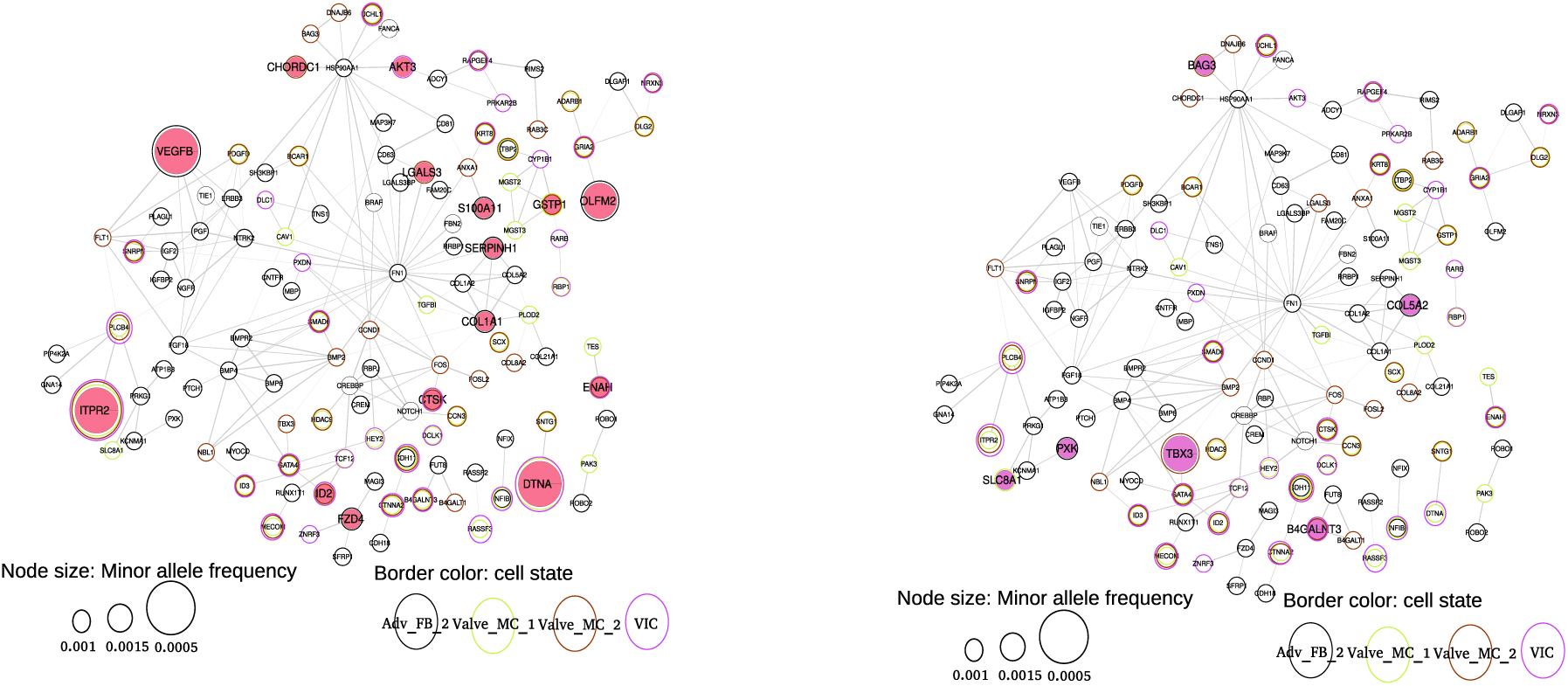
Single-patient networks of BAV2 and BAV3.

**Fig. S4:**
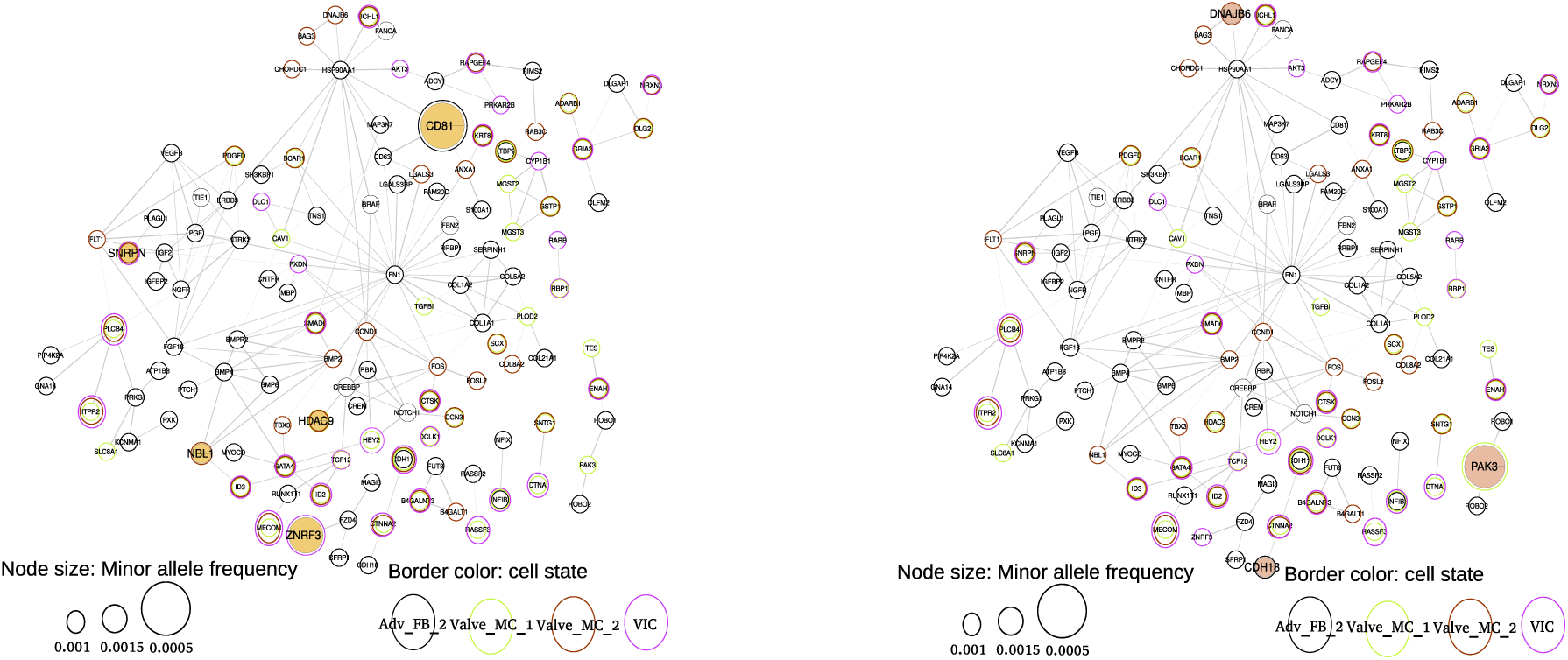
Single-patient networks of BAV5 and BAV6.

**Fig. S5:**
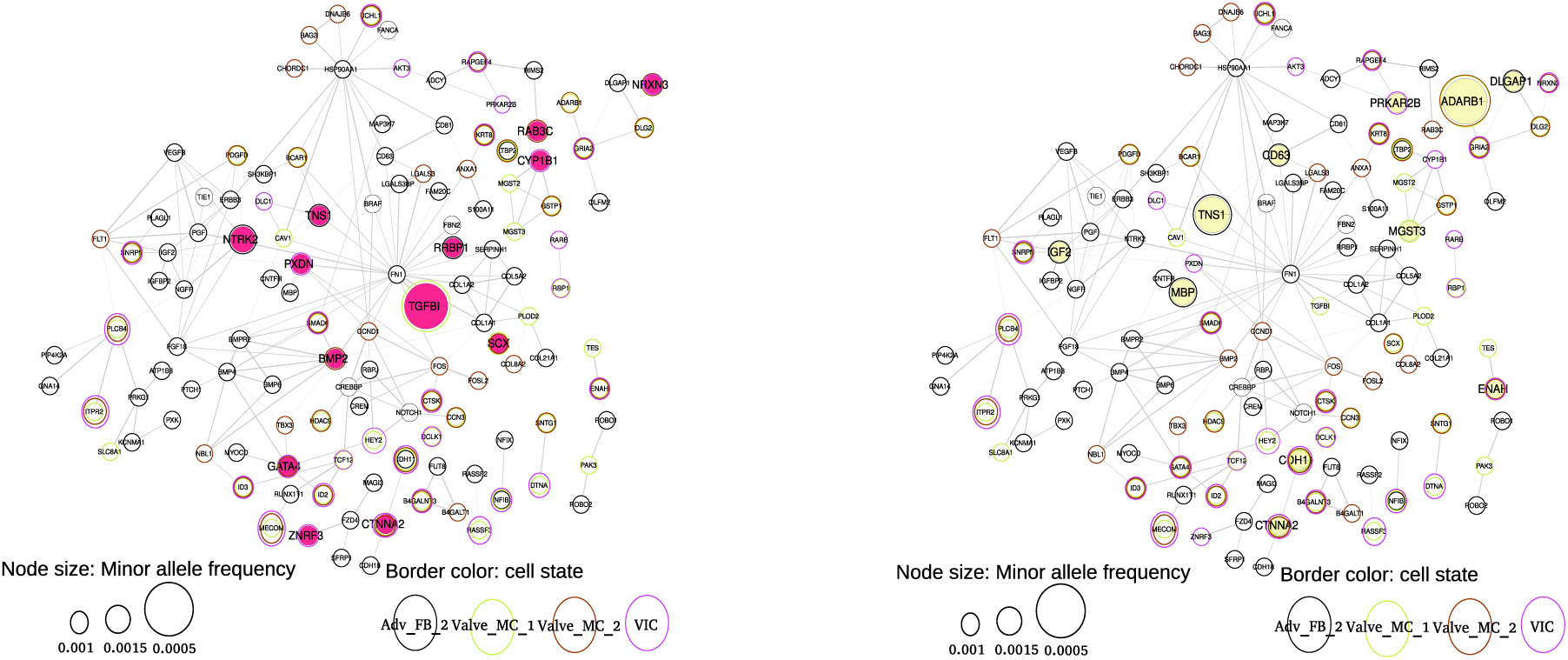
Single-patient networks of BAV7 and BAV8.

**Fig. S6:**
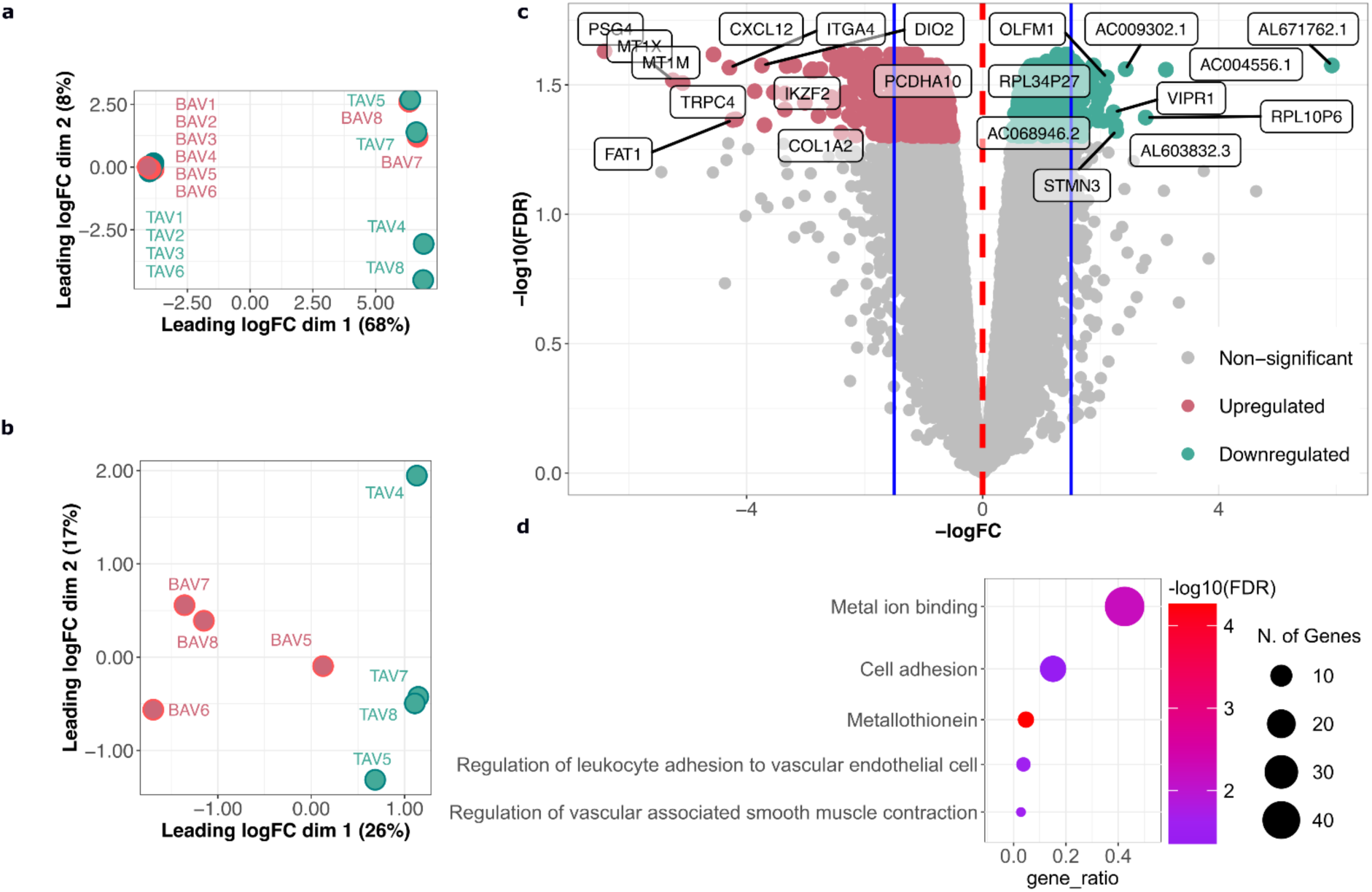
RNA-Seq analysis. **a,** MDS plot with all the patients. **b,** MDS plot upon removal of outliers. **c,** Volcano plot summarizing differentially expressed genes (DEGs) from bulk RNA-seq analysis. Grey dots represent non-significant DEGs. Green and red dots correspond to upregulated DEGs in TAV and BAV cohorts, respectively, with the top ten DEGs labeled by logFC. **d,** STRINGdb gene set enrichment analysis. Selected representative gene sets are displayed, illustrating significant pathways related to oxidative stress, metal ion binding, and inflammation in BAV patients.

