## Supplementary figures and images for "Rare gain-of-function regulatory mutations explain the missing heritability of bicuspid aortic valve"

### Figure S1

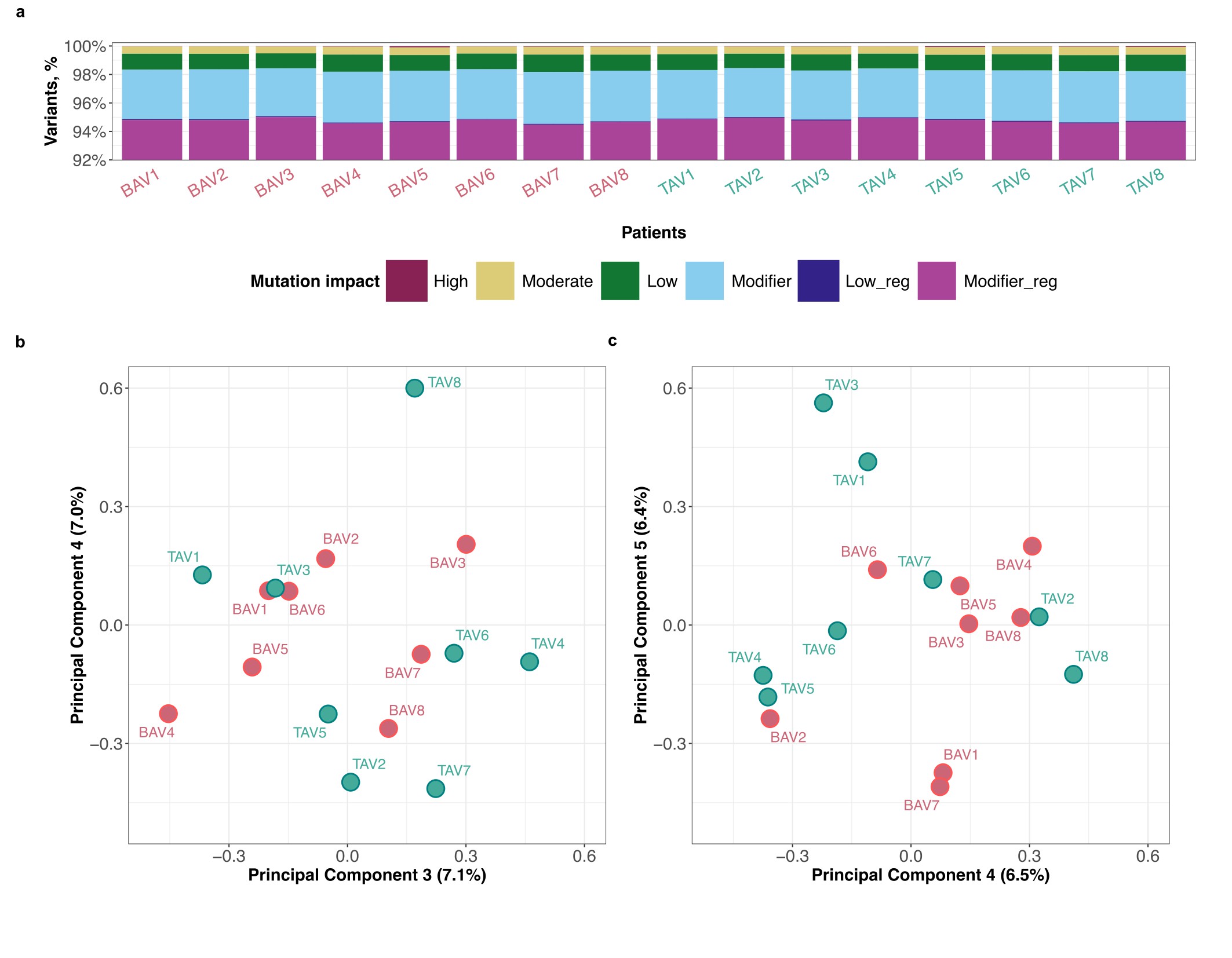

### Figure S2

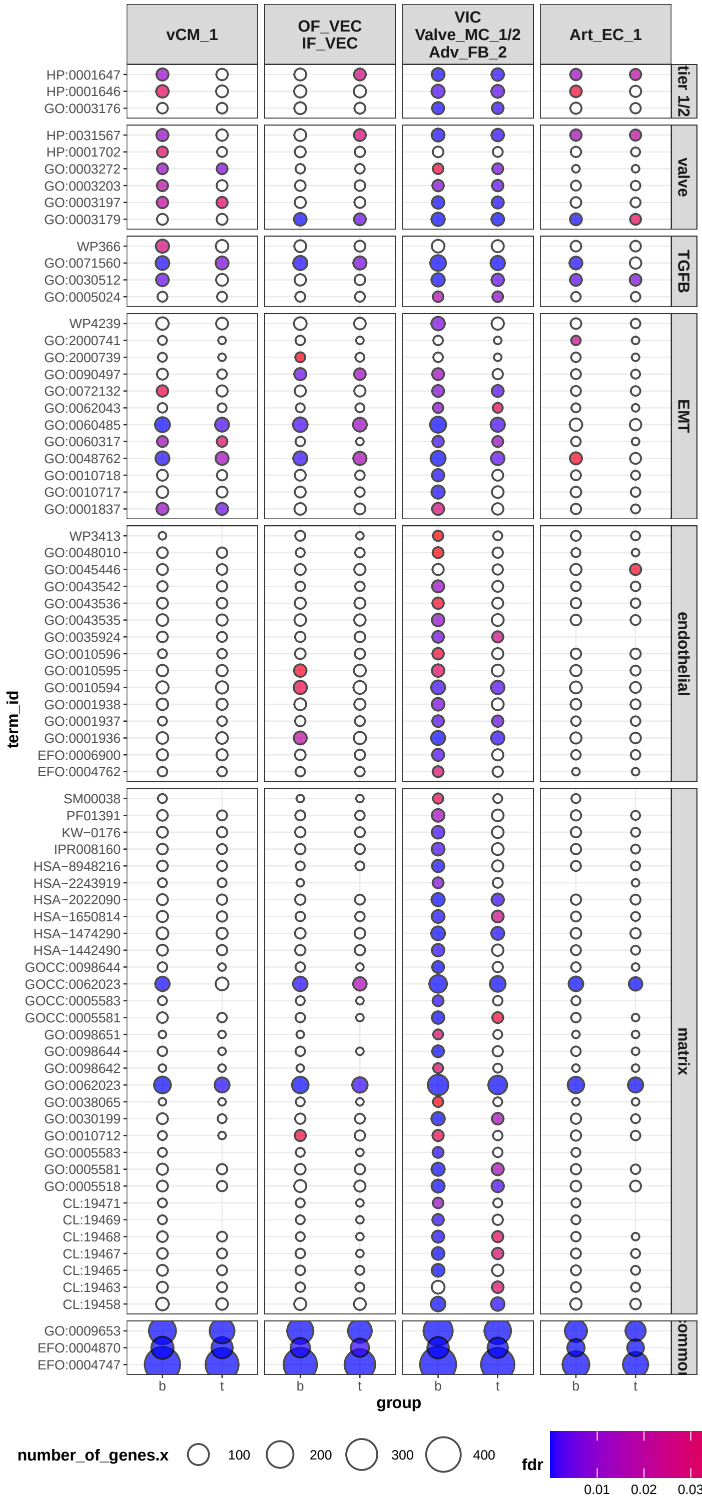

### Figure S3

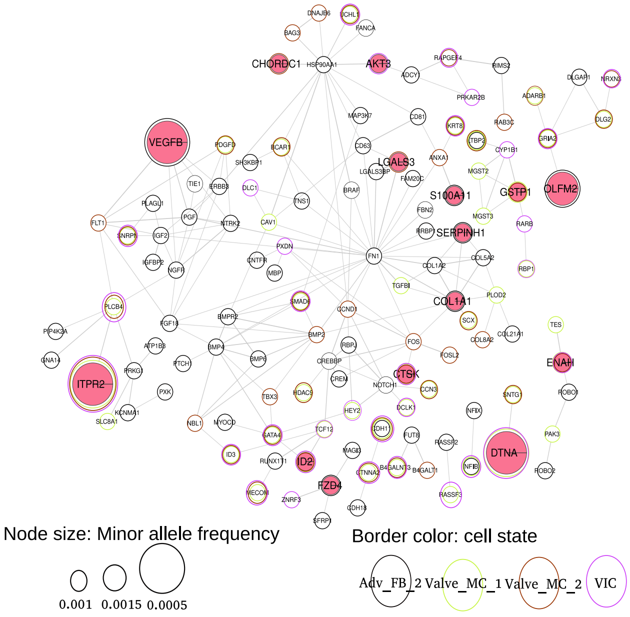

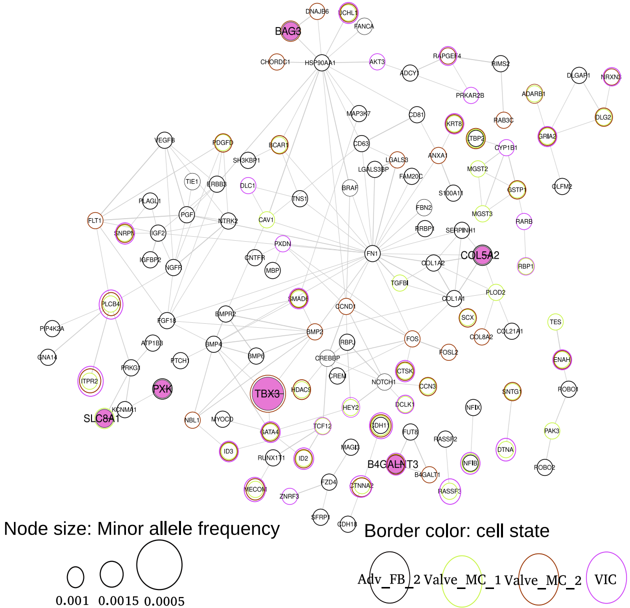

### Figure S4

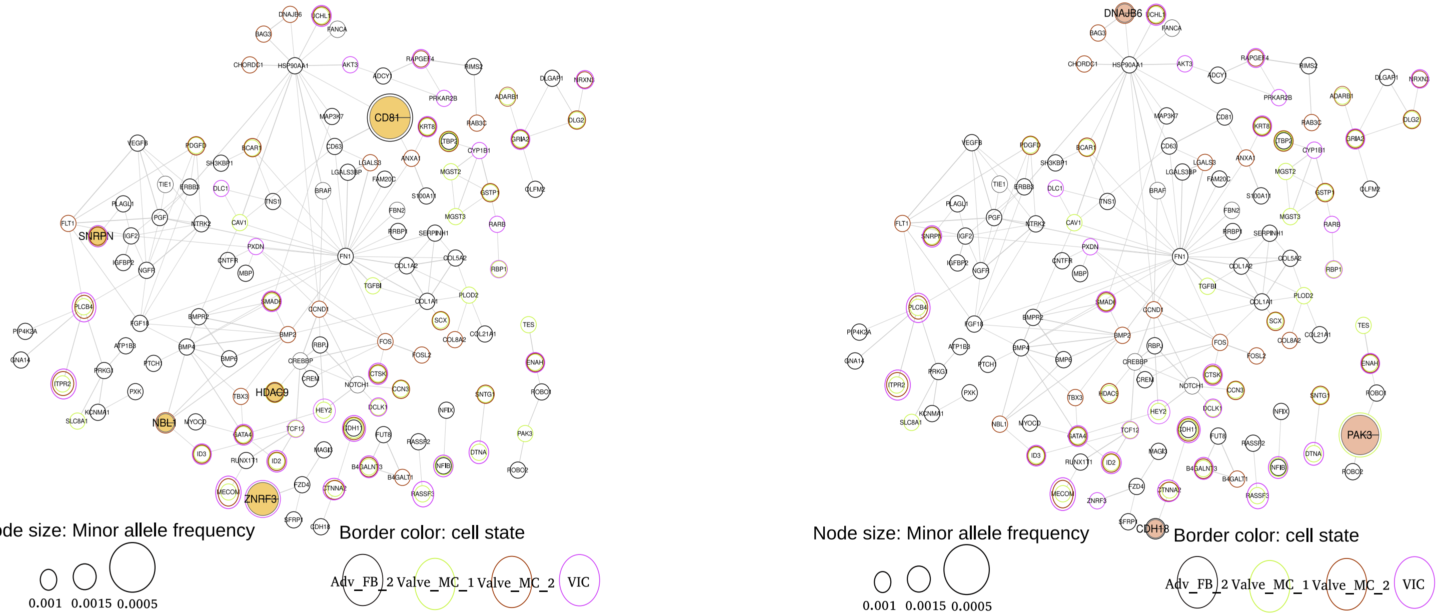

### Figure S5

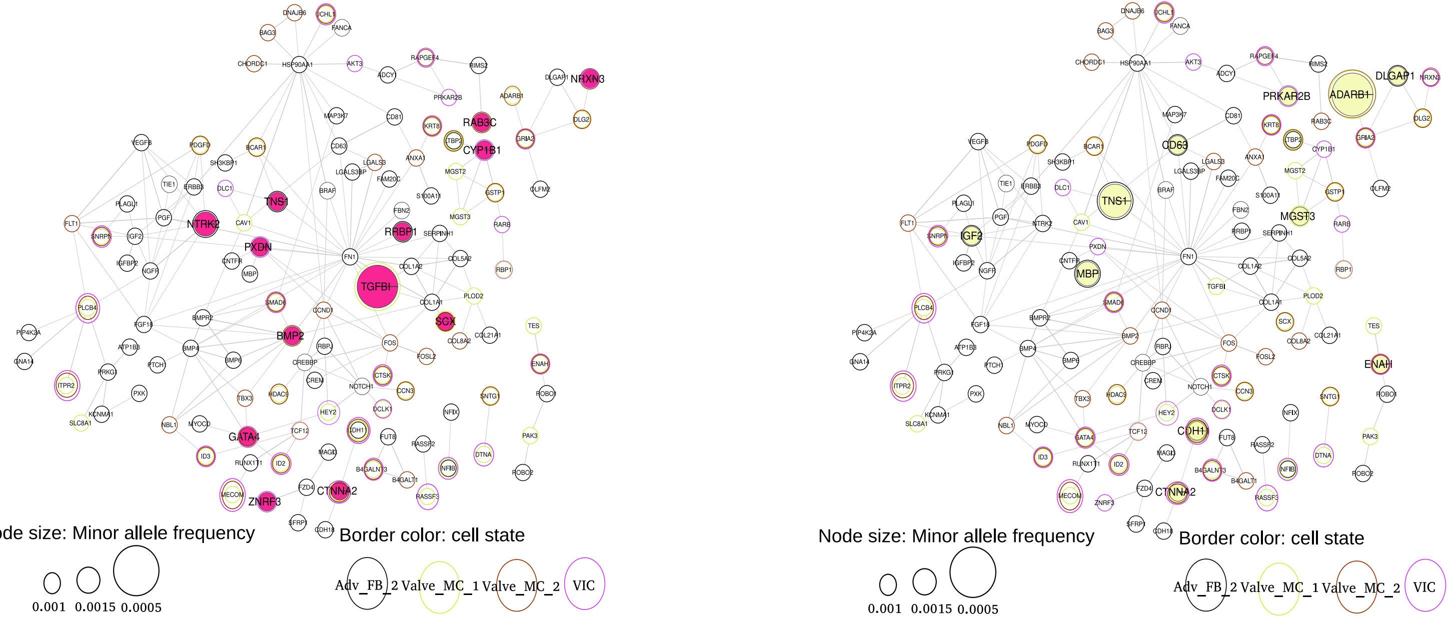

### Figure S6

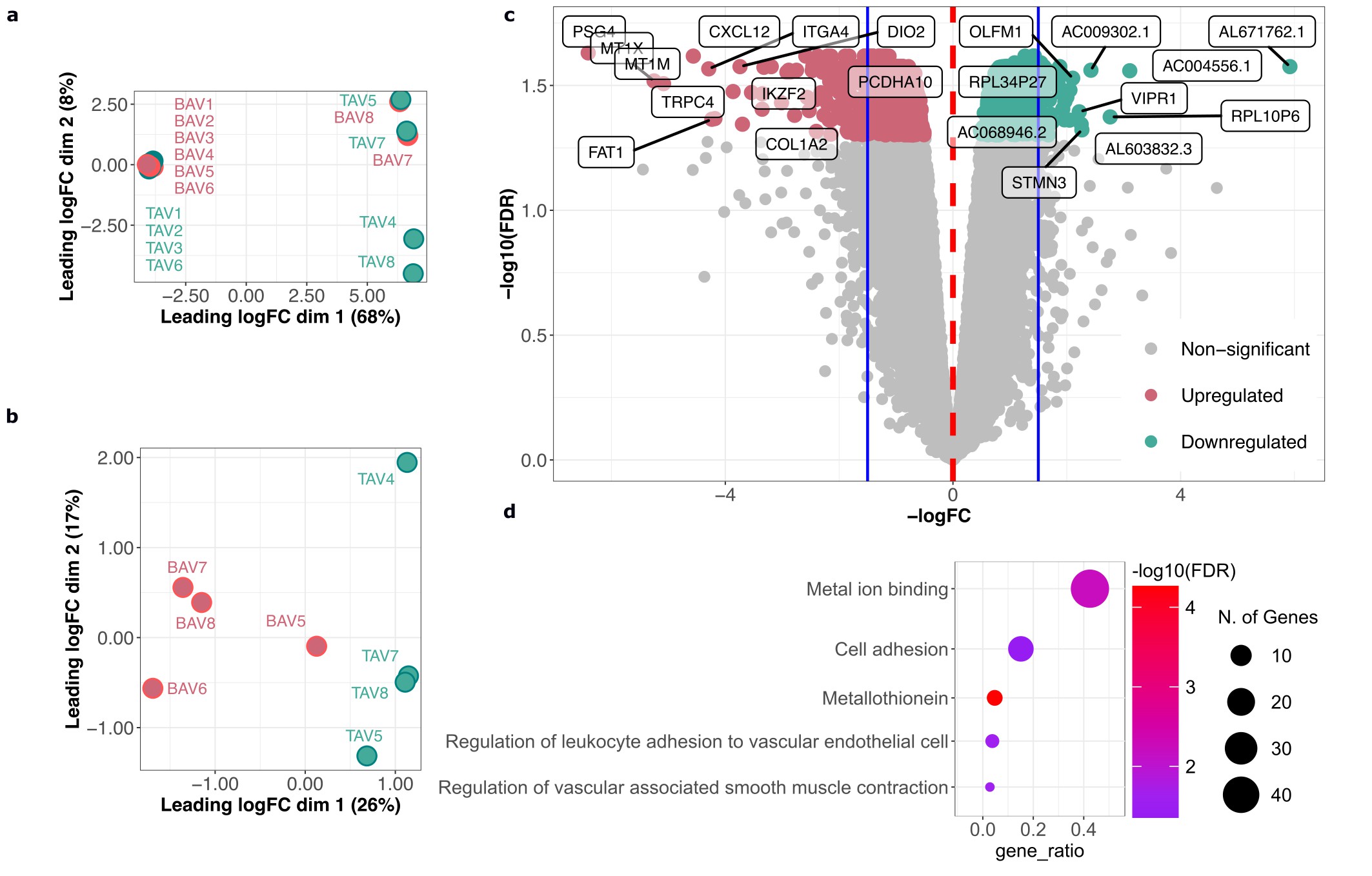
